# Development and external validation of a diagnostic multivariable prediction model for a prompt identification of cases at high risk for SARS-COV-2 infection among patients admitted to the emergency department

**DOI:** 10.1101/2021.05.26.21257834

**Authors:** Nicola Ughi, Antonella Adinolfi, Michel Chevallard, Laura Belloli, Michele Senatore, Alessandro Toscano, Andrea Bellone, Cristina Giannattasio, Paolo Tarsia, Massimo Puoti, Francesco Scaglione, Fabrizio Colombo, Michaela Bertuzzi, Giuseppe Bettoni, Davide Ferrazzi, Alessandro Maloberti, Armanda Dicuonzo, Francesca Del Gaudio, Claudio Rossetti, Oscar Massimiliano Epis, On behalf of the Niguarda COVID group

## Abstract

**Background:** An urgent need exists for an early detection of cases with a high-risk of SARS-CoV-2 infection, particularly in high-flow and -risk settings, such as emergency departments (EDs). The aim of this work is to develop and validate a predictive model for the evaluation of SARS-CoV-2 infection risk, with the rationale of using this tool to manage ED patients.

**Methods:** A retrospective study was performed by cross-sectionally reviewing the electronical case records of patients admitted to Niguarda Hospital or referred to its ED in the period 15 March to 24 April 2020.

Derivation sample was composed of non-random inpatients hospitalized on 24 April and admitted before 22 April 2020. Validation sample was composed of consecutive patients who visited the ED between 15 and 25 March 2020. The association between the dichotomic outcome and each predictor was explored by univariate analysis with logistic regression models.

**Results:** A total of 113 patients in the derivation sample and 419 in the validation sample were analyzed. History of fever, elder age and low oxygen saturation showed to be significant predictors of SARS-CoV-2 infection. The neutrophil count improves the discriminative ability of the model, even if its calibration and usefulness in terms of diagnosis is unclear.

**Conclusion:** The discriminatory ability of the identified models makes the overall performance suboptimal; their implementation to calculate the individual risk of infection should not be used without additional investigations. However, they could be useful to evaluate the spatial allocation of patients while awaiting the result of the nasopharyngeal swab.

**Key Messages box:** *What is already known on this topic:* 1 year after the onset of the coronavirus disease 2019 (COVID-19) pandemic, the trend of its spread has not shown a substantial global reduction. An urgent need exists for efficient early detection of cases with a high risk of SARS-CoV-2 infection and a number of diagnostic prediction models have been developed, but a few models were externally validated in high-flow and –risk settings, such as emergency departments (EDs).

*What this study adds:* This study develops and validate predictive models for the evaluation of SARS-CoV-2 infection risk, with the rationale of using these tools to promptly manage patients who are afferent to the ED, allocating them accordingly to the risk of infection while awaiting swab result. History of fever, older age and low oxygen saturation showed to be significant predictors of the presence of SARS-CoV-2 infection. The use of laboratory findings, such as neutrophil count, showed to improve the discriminative ability of the model, even if its calibration and usefulness in terms of diagnosis is unclear.

## INTRODUCTION

1 year after the onset of the coronavirus disease 2019 (COVID-19) pandemic, the trend of its spread has not shown a substantial global reduction, with 238,031 new cases registered on 11 March 2021, 117,573,007 total confirmed cases and 2,610,925 total deaths [1] and recurring new peaks of contagion are still observed locally in various geographical areas, including Italy (22,409 new cases registered on 11 March 2021, 3,123,368 total confirmed cases, 100,811 total deaths [2]).

Pending the effects of the vaccine, test and tracing methods still represent the most effective tools for containing the SARS-CoV-2 infection and preventing the spread of COVID-19 [3]. Rapid identification of SARS-CoV-2 infection is important for a prompt treatment and implementation of isolation procedures, particularly in high-flow and high-risk settings, such as emergency departments (EDs).

The standard reference test is represented by the search for the SARS-CoV-2 genome by reverse transcriptase-polymerase chain reaction (RT-PCR) of nasopharyngeal swab material, but although the technologies and execution times have been made more efficient, RT-PCR testing is time-consuming and its execution time (up to 4 hours) is often not compatible with urgent/emergency situations, consequently shorter decision times for the allocation of patients are still needed [4]. In order to reduce the identification time of SARS-CoV-2 cases, numerous models have been developed for the diagnosis of infection and disease, but with numerous concerns in terms of risk of bias. Consequently, the search for an improved predictive diagnostic tool is still open [5]. The aim of this work is to develop and validate a predictive model for the evaluation of SARS-CoV-2 infection risk, with the rationale of using this tool to promptly manage patients who are afferent to the ED, allocating them accordingly to the risk of infection while awaiting swab result.

## PATIENTS AND METHODS

### Study design and participants

A retrospective study was performed by cross-sectionally reviewing the electronical case records of patients who were admitted to Niguarda Hospital or referred to its ED after the first Italian COVID-19 case, between 15 March and 24 April 2020.

A non-random sample of inpatients from low-intensity general and specialist medical units who were hospitalized on 24 April 2020 and admitted before 22 April 2020, was used to develop the prediction model (derivation sample) and a sample of consecutive patients who visited the ED between 15 and 25 March 2020 was used to perform the historical external validation (validation sample).

Adult patients (≥18 years old) were eligible if at least one nasopharyngeal swab to search for the SARS-CoV-2 genome was performed, independently of the presence of the respiratory disease. The exclusion criteria were the following: current pregnancy, admission to pediatrics, obstetrics, surgery, psychiatric departments, or the high-intensity units, including critical care, at the time of inclusion.

All inpatients were tested for SARS-CoV-2 infection before the admission, while patients visiting ED were tested in the judgment of the attending physician. All the swab tests were processed at Niguarda Hospital.

The study is conformed to Helsinki’s Declaration and was approved by the ethics committee Milano Area 3 (register number 249-13052020). An informed consent was provided by the enrolled participants.

### Outcome and predictor measurements

The SARS-CoV-2 infection as the outcome was defined if at least one out of three SARS-Cov-2 genes tested on at least one nasopharyngeal swab was detected by RT-PCR.

For the derivation sample, the inpatients’ medical records were completed by interviews using *ad-hoc* case report forms to collect data about age, gender, communal living situations, cohousing and/or residence in care facilities, known contact with COVID-19 case; the presence (ever) of fever, cough, dyspnea, pharyngodynia, hypo-/anosmia and/or hypo-/ageusia, nausea and/or vomit, diarrhea; current history of syncope, seizure (if unprecedented), and/or conjunctivitis; systolic and diastolic blood pressures, heart and respiratory rates per minute, body temperature (°C), peripheral oxygen saturation while breathing ambient air; the total leukocyte, neutrophil, lymphocyte, and monocyte counts, and C-reactive protein levels (mg/dL, normal reference <0.5). The patient’s current medical history, symptoms and signs, vital signs, and laboratory exams at the time of the ED admission were considered as predictors.

The data needed for the validation sample were extracted by three physicians who reviewed the electronical medical records of the ED visits.

Data collection was unblinded to the outcome in both samples.

### Model development and validation methods

The association between the dichotomic outcome and each predictor was explored by univariate analysis with logistic regression models. The distribution of non-continuous predictors was checked and when the absolute frequency was less than 5 per table, they were not included in the model development due to potential separation and overfitting issues. Only main fixed effects were considered for inclusion in the model while interactions terms as well as higher order polynomials were not explored. A backwards stepwise algorithm based on the Akaike’s information criterion was used with bootstrap resampling (B = 1000) and the covariates whose statistical significance was detected in more than 80% of the bootstrap samples were chosen. The candidate models were compared in terms of performance by C-statistic, explained variation (McFadden’s R^2^; scaled) Brier score, Hosmer-Lemeshow test (grouping the observed outcomes by decile of predictions), and sensitivity and specificity (the Youden’s J statistic was used to identify the cutpoint for their maximization). The net reclassification improvement [6] was considered to further assess the effect of adding new predictors and the Decision Curve Analysis [7] was performed to evaluate the thresholds, as well as the quality of the predictions. Then, these models were internally validated by bootstrapping (B = 1000) [8] for estimation of the optimism-corrected C-statistic and calibration (slope and intercept). The original β coefficients of the candidate logistic models were used to calculate the predicted probabilities in an historical sample to assess their external validation and the above-mentioned performance measures were used. To explore the impact of overfitting due to small samples, a sensitivity analyses was performed by using shrunk [9] and penalized [10] estimations of the β coefficients and these predicted probabilities were compared.

Missing data were firstly managed by exploring the absence of remarkable differences between the complete cases and the group with missing data. Then, after checking the arbitrary-pattern of missing data and testing for the assumption of missing completely at random (Little’s chi-squared test) a complete case analysis was chosen both for the derivation and the validation samples.

The power to detect a statistically significant difference in the performance of the model at a particular sample size was calculated with standard formulas based on the normal distribution. The estimated size of the validation sample for a one-sample two-sided mean test was 351 to achieve 80% power if the mean difference between C-statistics 0.03 with standard deviation 0.20, and the type I error 0.05 were assumed.

All the analyses were performed using Stata Statistical Software Release 15 (StataCorp. 2017, College Station, TX: StataCorp LLC) and R (R Core Team 2018, R Foundation for Statistical Computing, Vienna, Austria).

The TRIPOD checklist for transparent reporting of a multivariable prediction model for individual prognosis or diagnosis was used as guidance for reporting the final version of this article [11]. The PROBAST tool was considered to discuss the potential risk of bias and concerns regarding model development and applicability [12].

## RESULTS

### Study population

A total of 576 patients were included in this study: 141 in the derivation sample and 435 in the validation sample. Of them, 28 patients in the derivation sample and 16 patients in the validation sample were excluded because of missing data, consequently 113 and 419 patients were analyzed, respectively (Figure 1).

**Figure 1.**
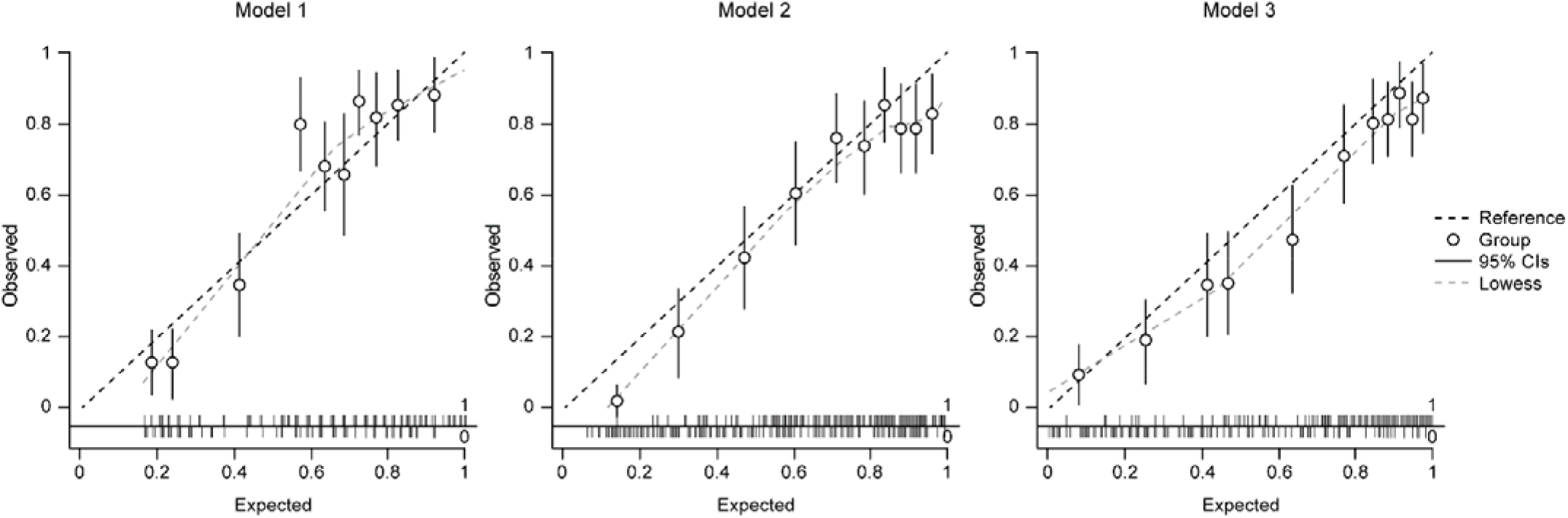
Calibration plots of the three models with the original coefficients in the external validation. CI: Confidence Interval.

The baseline characteristics of included patients are reported in Table 1.

**Table 1.** Baseline characteristics of included patients and case-mix features.

| **Features at presentation** | **Development sample**<br>**(n=141)** | **Validation sample**<br>**(n=435)** | **p-value** |
| --- | --- | --- | --- |
| Age (years), mean (SD) [range] | 72 (14) [27–96] | 61 (16) [19–93] | <0.01 |
| Male | 73 (51.8) | 267 (61.4) | 0.044 |
| Nasopharyngeal swab SARS-Cov-2 positivity | 71 (50.3) | 260 (59.8) | 0.047 |
| Report of contact with a person diagnosed of SARS-Cov-2 infection | 17/127 (13.4) | – | – |
| Communal living | 9/135 (6.7) | – | – |
| Hospitalization | 141 (100) | 373 (85.7) | <0.01 |
| Fever | 67/139 (48.2) | 300 (69.0) | <0.01 |
| Dyspnea | 42/139 (30.2) | – | – |
| Cough | 42/139 (30.2) | – | – |
| Pharyngodynia | 1/139 (0.7) | – | – |
| Hypo/anosmia | 1/139 (0.7) | – | – |
| Dysgeusia | 4/139 (2.9) | – | – |
| Conjunctivitis | 0/139 (0) | – | – |
| Nausea and/or vomit | 18/139 (12.9) | – | – |
| Diarrhea | 11/139 (7.9) | – | – |
| Seizure | 4/139 (2.9) | – | – |
| Syncope | 14/139 (10.1) | – | – |
| Body temperature (°C) | 36.6 (1.0) [35–39.8]<sup>^</sup> | – | – |
| Oxygen saturation (room air), % | 94.4 (4.2) [78–100]* | 93.6 (6.1) [40–100]<sup>†</sup> | 0.148 |
| Systolic blood pressure (mmHg) | 140 (31) [70–230]* | – | – |
| Diastolic blood pressure (mmHg) | 73 (14) [40–123]* | – | – |
| Heart rate per minute | 91 (20) [45–70]<sup>§</sup> | – | – |
| Respiratory rate per minute | 21 (6) [12–40]<sup>#</sup> | – | – |
| White blood cell count (10<sup>9</sup>/L) | 9.8 (6.7) [1.4–51.2] | – | – |
| Neutrophil count (10<sup>9</sup>/L) | 7.3 (5.3) [1.1–42.5]<sup>°</sup> | 6.4 (4.4) [0.7–46.2]<sup>‡</sup> | 0.046 |
| Lymphocyte count (10<sup>9</sup>/L) | 1.6 (2.1) [0.1–16.8]<sup>°</sup> | – | – |
| Monocyte count (10<sup>9</sup>/L) | 0.8 (1.5) [0.04–16.5]<sup>°</sup> | – | – |
| C-reactive protein (mg/dL) | 6.8 (7.9) [0–43.4] | – | – |
| Data presented as n (%) and mean (SD) [range].<br><sup>^</sup>4/141 (2.8%) missing; <sup>*</sup>2/141 (1.4%) missing; <sup>§</sup>1/141 (0.7%) missing; <sup>#</sup>24/141 (8.3%) missing;<br><sup>°</sup>3/141 (2.1%) missing; <sup>†</sup>2/435 (0.5%) missing; <sup>‡</sup>15/435 (3.4%) missing.<br>SD, standard deviation |  |  |  |

Regarding the case-mix, a significant higher proportion of positive outcome (71 out of 141, 50% vs 260 out of 435, 60%; p=0.047), male patients (73 out of 141, 52% vs 267 out of 435, 61%, p=0.044) with fever (67 out of 139, 48% vs 300 out of 435, 69%, p≤0.01) were observed in the validation sample compared to the derivation sample, while in the same sample age and neutrophil counts were lower (respectively, mean [SD] age equal to 72 [14] vs 61 [16] years, p <0.01, and neutrophil mean [SD] count equal to 7.3 [5.3] vs 6.4 [4.4] 10^9^ cells/L) (Table 1).

The differences in terms of outcome and predictors between the patients analysed and those excluded were not statistically significant with the exception of a higher prevalence of cough in the complete cases (39 out of 113, 34% vs 3 out of 26, 11%, p=0.03) of the derivation sample and a lower frequency of hospitalization (363 out of 419, 87% vs 10 out of 16, 62%, p≤0.01) in the validation set (Supplementary Table 1).

The average time between the patient’s assessment and the test result of swab was less than 24 hours.

### Prediction models

In the derivation sample, age, oxygen saturation at room air, fever, cough, and both total white blood cell and neutrophil counts were significantly associated in the univariate analyses (Table 2). In the selection of the predictors, only age, oxygen saturation at room air, fever, and the neutrophil count (total white blood cell was excluded due to high correlation with the individual cell count) showed a statistical significance in more than 80% of the 1000 bootstrap samples and they were considered for model building.

**Table 2.** Predictors included in the selection process and results of their univariate analyses in the two study samples.

|  |  |  |
| --- | --- | --- |
|  | **OR (95% CI)** |  |
|  | **Derivation sample**<br>**(n=113)** | **Validation sample**<br>**(n=419)** |
| Age, 1-year increase | 0.95 (0.93-0.99) | 0.99 (0.97-1.00) |
| Male, female as reference | 1.19 (0.57-2.50) | 1.21 (0.81-1.81) |
| Fever, absence as reference | 5.88 (2.61-13.23) | 18.64 (10.87-31.98) |
| Dyspnoea, absence as reference | 1.59 (0.72-3.55) | — |
| Cough, absence as reference | 3.3 (1.45-7.52) | — |
| Nausea and/or vomit, absence as reference | 0.71 (0.23-2.18) | — |
| Syncope, absence as reference | 0.63 (0.17-2.36) | — |
| Body temperature (°C) | 1.44 (0.95-2.13) | — |
| Oxygen saturation (room air), 1% increase | 0.86 (0.78-0.96) | 0.86 (0.81-0.90) |
| Systolic blood pressure, 10-mmHg increase | 1.02 (0.91-1.14) | — |
| Diastolic blood pressure, 10-mmHg increase | 1.07 (0.84-1.37) | — |
| Heart rate, 10-beat-per-minute increase | 1.07 (0.89-1.30) | — |
| Respiratory rate, 1-breath-per-minute increase | 1.01 (0.95-1.07) | — |
| White blood cell count, $1 \times 10^9/L$ increase | 0.86 (0.79-0.95) | — |
| Neutrophil count, $1 \times 10^9/L$<br>increase | 0.87 (0.78-0.96) | 0.88 (0.83-0.93) |
| Lymphocyte count, $1 \times 10^9/L$<br>increase | 0.61 (0.37-1.02) | — |
| Monocyte count, $1 \times 10^9/L$<br>increase | 0.70 (0.39-1.26) | — |
| C-reactive protein, 1-mg/dL<br>increase | 0.96 (0.92-1.01) | — |
| OR, odds ratio; CI, confidence interval. |  |  |

Three multivariable models were consequently developed: an oversimplified one with fever and oxygen saturation only (M1), and two models with the clinical predictors in absence (M2) and presence (M3) of the results of the laboratory test, and their apparent performance was tested as reported in Table 3.

**Table 3.**
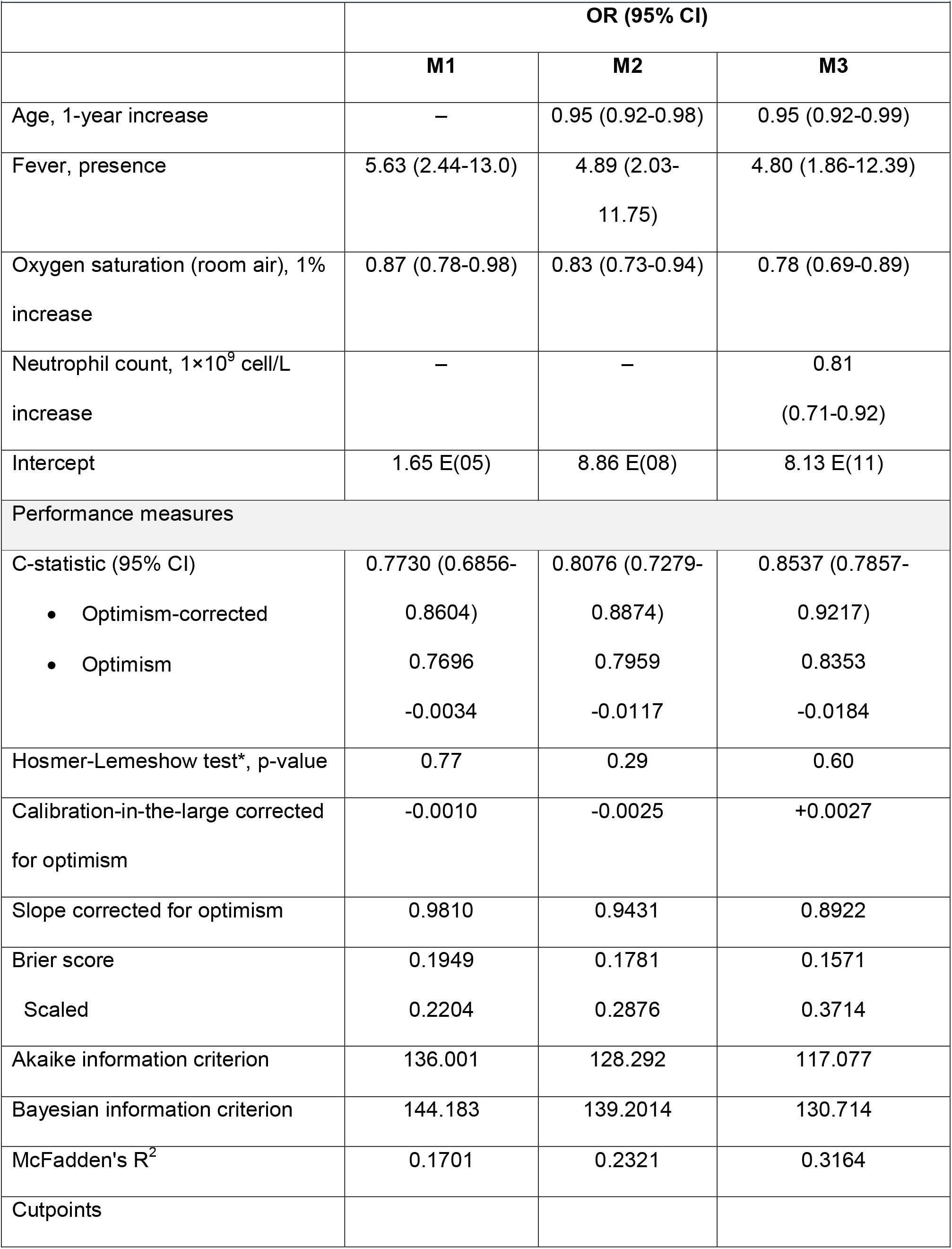
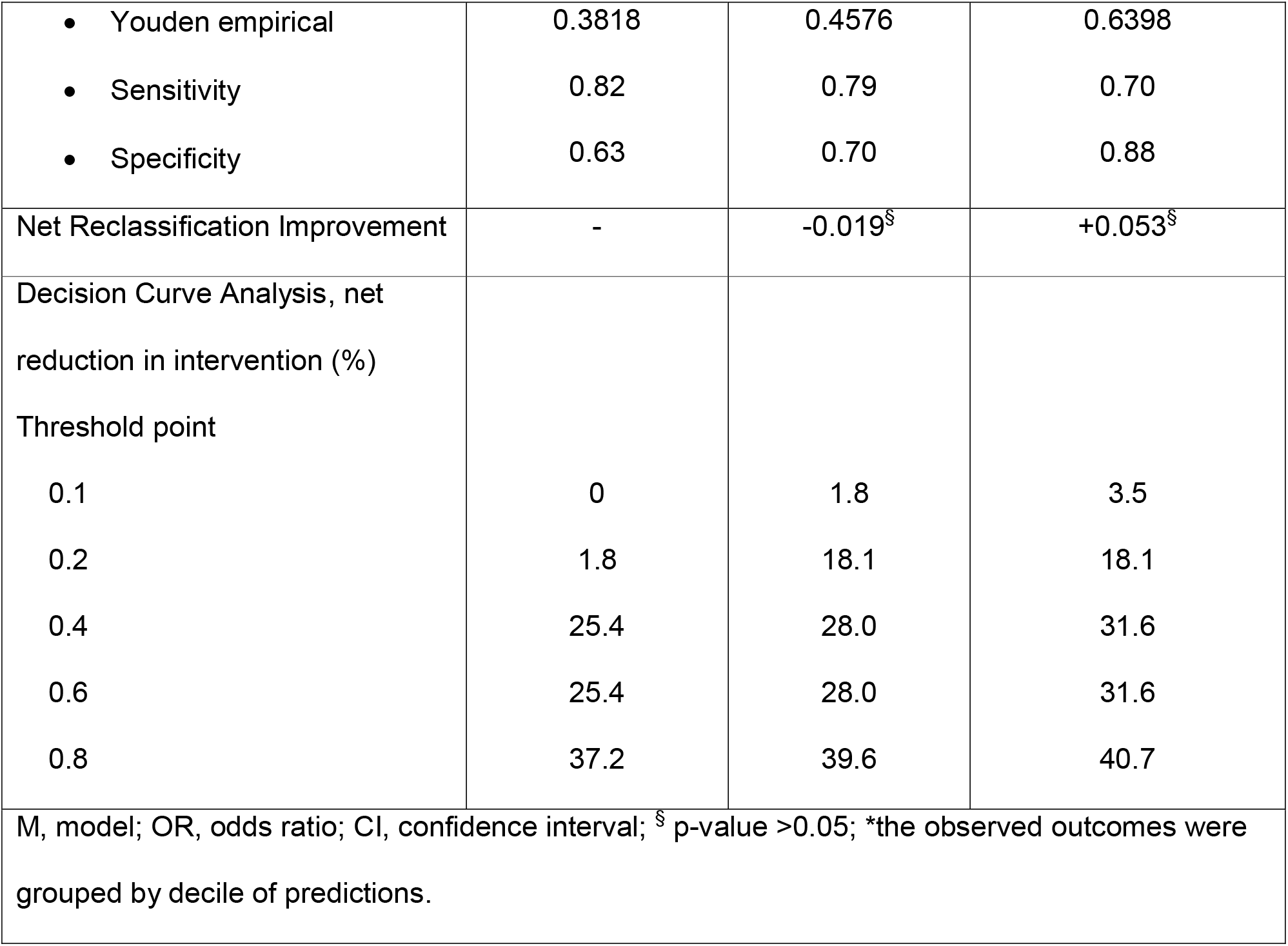
Performance of the three multivariable models in the development sample (n=113)

The discrimination was >0.80 in M2 and M3, but the net reclassification improvement due to the addition of the neutrophil count as predictor was not statistically significant. In the view of Decision Curve Analysis, M2 was steadily superior to M1, and inferior to M3, but not for all the threshold points. When these models were internally validated, the correction of the calibration slope for optimism was <10% in M1 and M2 only.

### Validation results

The external validation of the models and the sensitivity analyses of the regression coefficients by uniform shrinking and penalization are detailed in Table 4 and Supplementary Table 2.

**Table 4.**
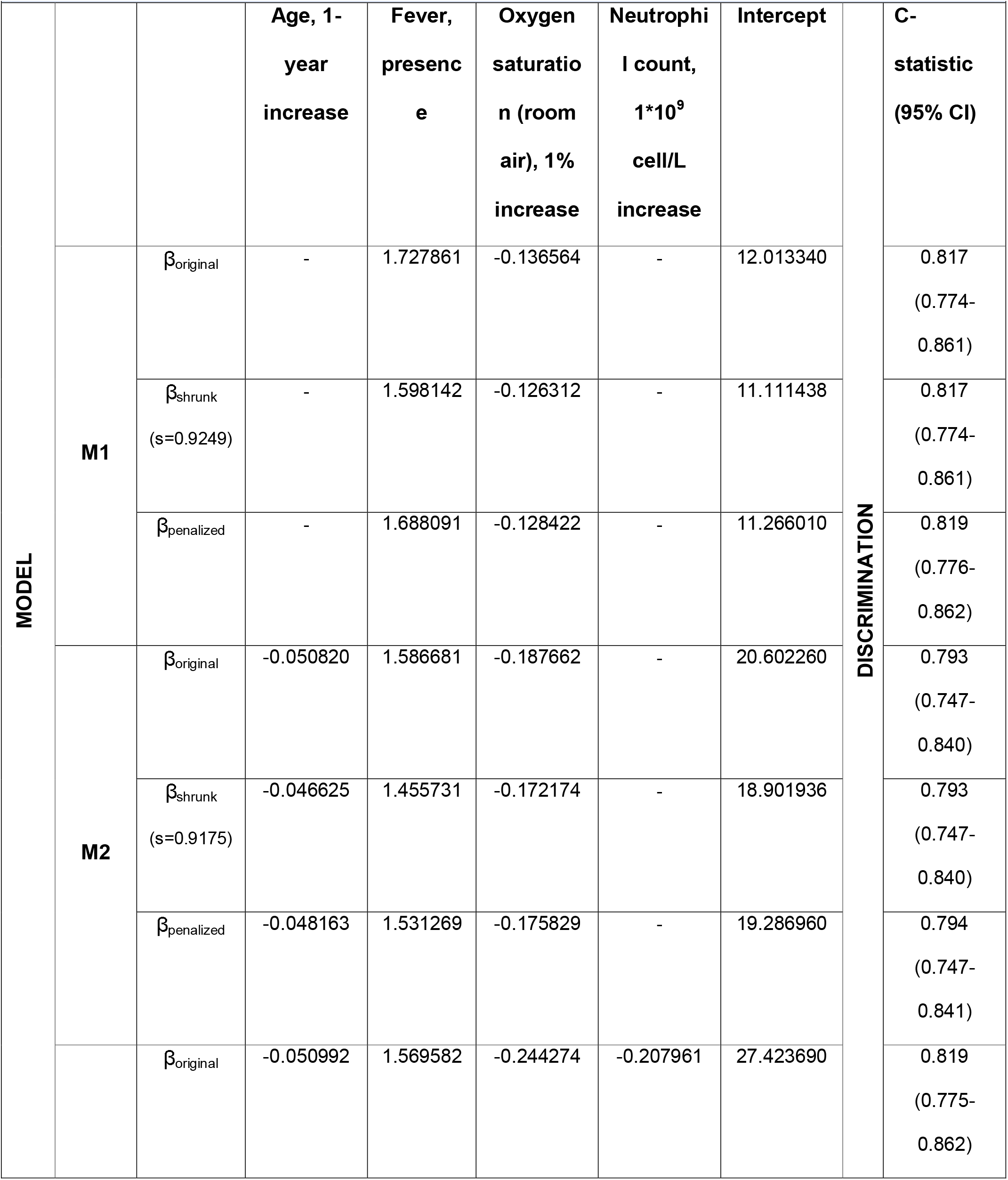
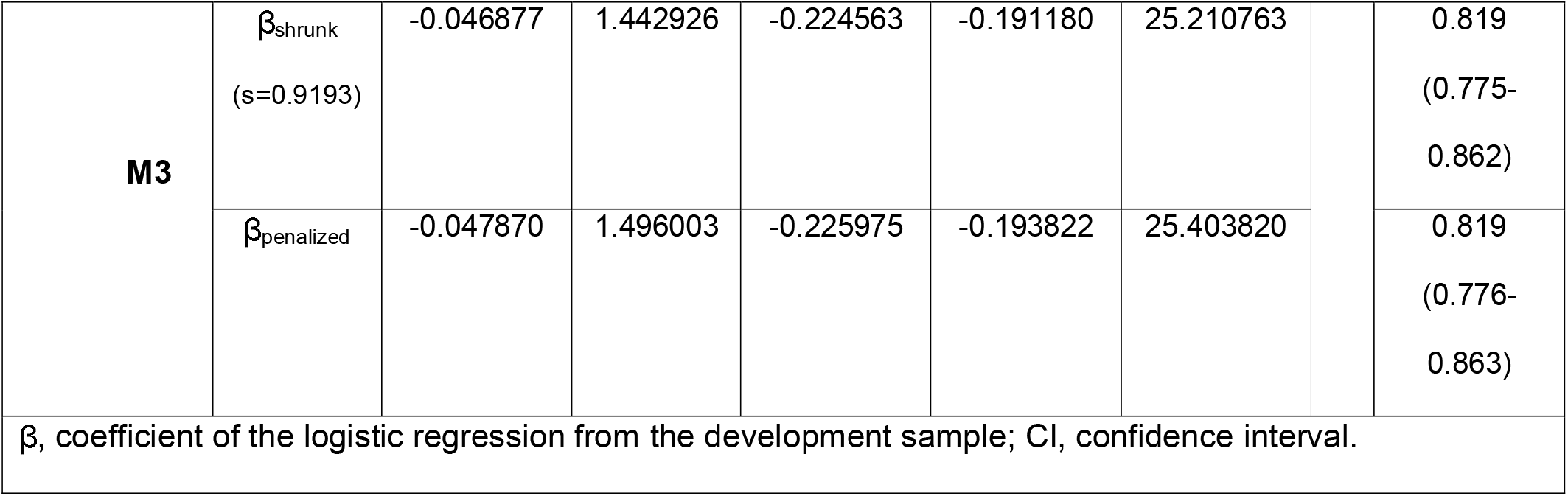
Intercept and coefficients of the three diagnostic models (original and values modified by uniform shrinking and penalization) and their discrimination performance in the validation sample (n=419).

The C-statistic was significantly higher for M3 (0.819, 95%CI 0.775-0.862) compared to M2 (0.793, 95% 0.747-0.840, p=0.014), yet the difference was limited (2.6%) and statistically not significant in comparison with M1 (0.817, 95% CI 0.774-0.861, p=0.933). However, only M2 showed to be sufficiently well calibrated if shrunk or penalized coefficients were applied instead of the original values. The calibration plots based on the original coefficients are shown in Figure 2. The sensitivity was consistently >85% for all the models, but the specificity was >70% in M3 only when the cut-off was statistically calculated to maximize both the performance measures (Supplementary Table 2). In Figure 3, the curves show that each model is clinically useful for thresholds in the range of 20-80% for the probability of the outcome. Moreover, each model showed to be superior to the use of age alone as well as the oxygen saturation at room air, while the history of fever as the only predictor seemed to be comparable.

**Figure 2.**
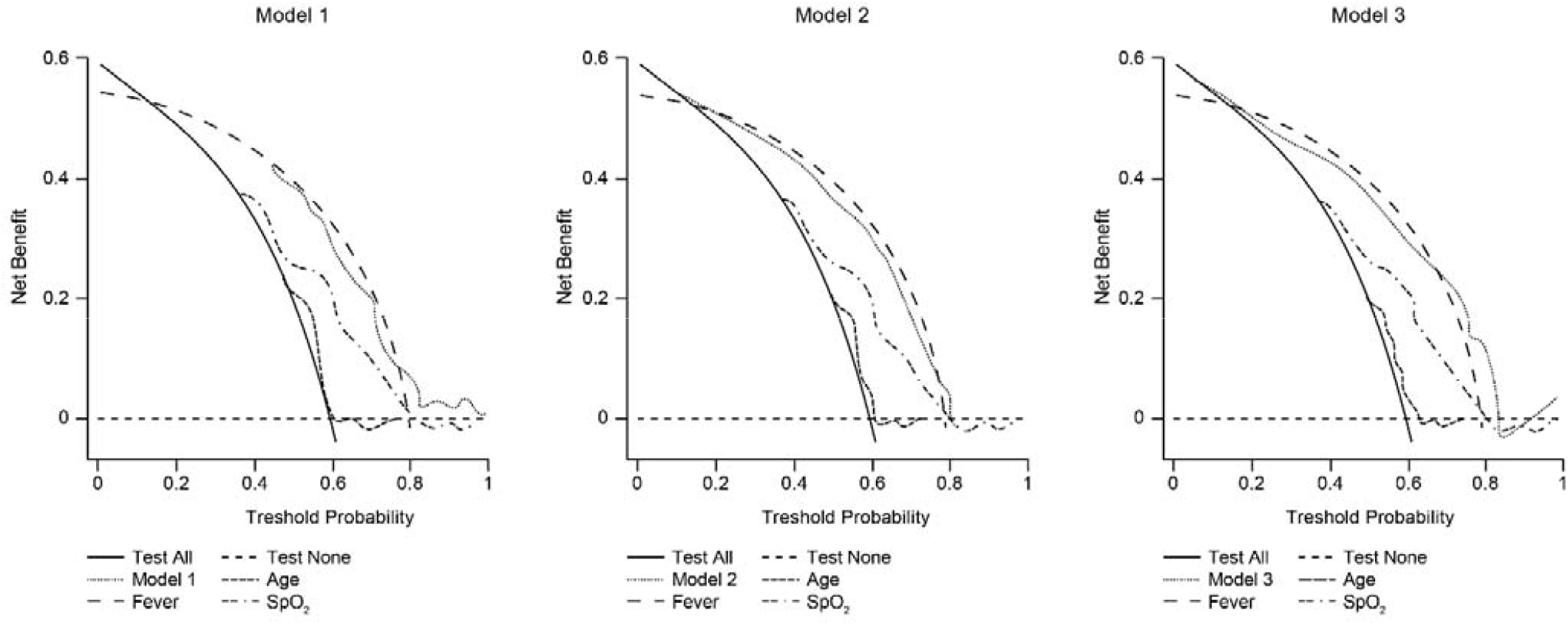
The Decision Curve Analysis of the three models applied into the validation sample in comparison with the use of the individual predictors across different probability thresholds for clinical decision. SpO_2_: oxygen saturation.

## DISCUSSION

Niguarda is one of the largest General Hospital in the North of Milan within a Metropolitan Area of 3,279,944 inhabitants (January 2020), and hosts all the medical and surgical disciplines for adults and children, including a 24-hour ED with 96,588 visits and 32,612 in hospital admissions covering every intensity of care in 2019.

In this hospital context, a prediction model for the prompt diagnosis of COVID-19 high risk was developed in hospitalized patients and externally validated in the setting of the ER.

The history of fever, elder age and low oxygen saturation showed to be significant predictors of the presence of SARS-CoV-2 infection in the derivation sample. The use of laboratory findings, such as neutrophil count showed to improve the discriminative ability of the model, even if its calibration and usefulness in terms of diagnosis is unclear.

Since the beginning of the pandemic, a remarkable number of prediction models have been developed and critically appraised in a systematic review updated to July 2020 [5]. Out of 232 models, 118 were diagnostic models for detecting COVID-19. Of them, 75 were based on medical imaging and 10 were used to diagnose disease severity. Only few diagnostic models were externally validated, and all the appraised models were judged to be flawed by a high risk of bias [5].

Age was included in most of the final models, while the inclusion of fever or varying laboratory findings, including neutrophil count, were inconsistent and the oxygen saturation was considered only in prognostic models (107 out of 232), despite its clinical importance in the early management of disease [5].

Among the prediction models appraised, the diagnostic model by Jehi et al. was identified as promising, even if requires external validation. It defines male, African–American, older patients, and those with known COVID-19 exposure at higher risk of being positive for COVID-19 [13]. A reduced risk was reported for patients who had influenza or pneumococcal polysaccharide vaccine or who were on melatonin, paroxetine or carvedilol. Nevertheless, the inclusion of race as predictor is subjected to a possible bias due to American healthcare inequality, and the COVID-19 exposure is a data reported by patients [13].

The role of social variables within a COVID-19 predictive model was explored by Chew and collaborators, who derived two models which include, among different risk factors, dormitory residence in the first and contact with infective patients in the second model [14].

Considering the dormitory residence, as the institutions dedicated to migrants, authors conclude that this represents an important predictor for COVID-19 infection, but this result is not confirmed if we consider other types of institutions, such as retirement homes. This result is therefore not generalizable to realities other than Singapore.

Considering contact with COVID-19 infective patients, authors highlight the role of traceability as prognostic factor in the development of the disease. On the other hand, contact tracing is not easy to execute, both in traditional and alternative ways, because of privacy and socio-cultural aspects [15].

Both considerations could explain the lack of predictability of residency data in some specific contexts and exposure data in the present study [14].

Of note, Italian studies are increasing accordingly to the last update of the systematic review which counts a total of 23 (10%) Italian models, 21 more than the first version of April 2020.

The discriminative ability of our models is consistent with those whose performance was assessed by external validation with C-statistics ranging from 0.73 to 0.91. Moreover, their calibration was superior to that of the only study where the calibration slope was reported (0.77 to 0.95 vs 0.56, respectively) [16].

With regards to the setting, the only study based on patients from ED reported a C-statistic calculated by training test split (0.85) comparable to those of our models in the derivation sample, but the number of predictors was higher (15 versus 2 to 4 variables) and its external validation was not provided, and this may raise concerns about overfitting issues [17]. Moreover, another prediction model for diagnosis in Italy was developed to assess the probability of community-acquired pneumonia due to COVID-19 from multiple centres and its discriminative ability was similar (C-statistic 0.84) [18]. However, this comparison may be flawed by the differences between the purposes of the models as well as their settings (hospitalized patients vs ED admissions). The implementation of our models is based on a limited number of predictors which are easy to be measured as well as promptly available in everyday practice. Moreover, its temporal external validation showed to be robust to the differences in the case-mix and particularly when the prevalence of the outcome and the proportion of hospitalization were variable in real-life consecutive sample of patients from the ED setting.

This study has several limitations. First, it was developed by using a case-control design and case analysis and data collection were retrospective and unblinded. However, since hard outcome and predictors like the detection of SARS-CoV-2 genome in the nasopharyngeal swab and age were used, a limited impact of bias on predictions may be assumed. Secondly, predictors like communal living and history of contact with a person diagnosed of SARS-CoV-2 infection were unmeasured, even though their relevance for diagnostic predictions has been still to be determined. Finally, the discriminatory ability of the models makes the overall performance still suboptimal and its implementation to calculate the individual risk of infection should not be used without additional investigations.

However, it could be considered to evaluate the spatial allocation of the patients while awaiting the result of the nasopharyngeal swab, which is still the current reference standard for the diagnosis of SARS-Cov-2 infection.

In conclusion, the development and validation of these models showed that prediction tools based on clinical findings like history of fever, age, and oxygen saturation at presentation may be accurate and robust to identify patients at high risk for a diagnosis of SARS-CoV-2 infection. Future external validation studies should be considered to evaluate if such models will be also robust to variable outcome prevalence, particularly in low proportions of infection, and to search for additional predictors which could increase the model performance for individual risk prediction.

## Data Availability

Data may be made available upon reasonable request.

## Supplementary Material

Supplementary Table 1.

Supplementary Table 2.

## Availability of data and material (data transparency)

Data may be made available upon reasonable request.

## Ethics approval

The study was conducted within the protocol approved the ethics committee Milano Area 3

(register number 249-13052020).

## Consent to participate

All the participants signed an informed consent form

## Consent for publication

Not required as this manuscript doesn’t include details, images or videos related to the participants

## Acknowledgments

Editorial assistance was provided by Simonetta Papa, PhD, and Aashni Shah (Polistudium SRL, Milan, Italy). Graphical assistance was provided by Massimiliano Pianta (Polistudium SRL, Milan, Italy).

