## Supplementary Material for "Development and external validation of a diagnostic multivariable prediction model for a prompt identification of cases at high risk for SARS-COV-2 infection among patients admitted to the emergency department"

| **Table S1. Comparisons between the patients included for the analyses and those excluded due to missing data both in the development and the validation sample.** | | | | | | |
| --- | --- | --- | --- | --- | --- | --- |
| **Features** | **Development sample (n=141)** | | **p-value** | **Validation sample (n=435)** | | **p-value** |
|  | **Complete cases, n=113 (80.1%)** | **Missing data group, n=28 (19.9%)** |  | **Complete cases, n=419 (96.3%)** | **Missing-data group, n=16 (3.7%)** |  |
| Age (years), mean (SD) [range] | 72 (14)  [27–96] | 72 (15) [48–96] | 0.890 | 61 (16)  [19–93] | 63 (12)  [38–89] | 0.789 |
| Male | 59 (52.1) | 14 (50.0) | 0.834 | 255 (60.9) | 12 (75.0) | 0.305 |
| Nasopharyngeal swab SARS-CoV-2 + | 57 (50.4) | 14 (50.0) | 0.967 | 252 (60.1) | 8 (50.0) | 0.417 |
| Patient’s report of contact with a person diagnosed with SARS-CoV-2 infection | 14/102 (13.7) | 3/25 (12.0) | 1.000 | – | – | – |
| Community residence | 7/107 (6.5) | 2/28 (7.1) | 1.000 | – | – | – |
| Hospitalization | 113 (100) | 28 (100) | – | 363 (86.6) | 10 (62.5) | 0.007 |
| Fever | 58  (51.3) | 9/26  (34.6) | 0.124 | 290 (69.2) | 10 (62.5) | 0.569 |
| Dyspnea | 36  (31.9) | 6/26  (23.1) | 0.379 | – | – | – |
| Cough | 39 (34.5) | 3/26 (11.5) | 0.031 | – | – | – |
| Pharyngodynia | 0 (0) | 1/26 (3.8) | 0.187 | – | – | – |
| Hypo/anosmia | 1 (0.9) | 0/26 (0) | 1.000 | – | – | – |
| Dysgeusia | 3 (2.6) | 1/26 (3.8) | 0.568 | – | – | – |
| Conjunctivitis | 0/113 (0) | 0/26 (0) | - | – | – | – |
| Nausea and/or vomit | 14 (12.4) | 4/26 (15.4) | 0.746 | – | – | – |
| Diarrhea | 9 (8.0) | 2/26 (7.7) | 1.000 | – | – | – |
| Seizure | 4 (3.5) | 0/26 (0) | 1.000 | – | – | – |
| Syncope | 10 (8.8) | 4/26 (15.4) | 0.297 | – | – | – |
| Body temperature (°C), mean (SD) [range] | 36.6 (1.0)  [35–39.8] | 36.5 (1.0)  [35–38.8] | 0.628 | – | – | – |
| Oxygen saturation (room air; %) | 94.2 (4.3)  [78-100] | 95.4 (3.8)  [80-100] | 0.183 | 93.5 (6.2)  [40-100] | 94.9 (4.4)  [84-100] | 0.432 |
| Systolic blood pressure (mmHg) | 139 (32)  [70–230] | 144 (30)  [95–210] | 0.448 | – | – | – |
| Diastolic blood pressure (mmHg) | 73 (15)  [40–123] | 78 (12)  [55–100] | 0.164 | – | – | – |
| Heart rate per minute | 91 (20)  [45–158] | 91 (23)  [60–170] | 0.978 | – | – | – |
| Respiratory rate per minute | 21 (6)  [12–40] | 23 (6)  [18–30] | 0.558 | – | – | – |
| White blood cell count (10^9^/L) | 9.8 (7.1)  [2.4–51.2] | 9.6 (5.4)  [1.4–22.4] | 0.887 | – | – | – |
| Neutrophil count (10^9^/L) | 7.5 (5.4)  [1.2–42.5] | 6.4 (4.2)  [1.1–17.4] | 0.361 | 6.4 (4.4)  [0.7-46.2] | 18.3 (–)  [–] | – |
| Lymphocyte count (10^9^/L) | 1.6 (1.8)  [0.1–15.9] | 1.8 (3.2)  [1.1–17.4] | 0.559 | – | – | – |
| Monocyte count (10^9^/L) | 0.9 (1.6)  [0.05-16.5] | 0.6 (0.4)  [0.04-1.5] | 0.402 | – | – | – |
| C-reactive protein, mg/dL | 7.3 (8.1)  [0-43.4] | 4.9 (6.5)  [0.1–26.9] | 0.170 | – | – | – |
| Data are presented as n (%) or mean (SD) [range]. SD, standard deviation; P values were calculated by two-sided two-sample t tests or two-sided Pearson chi-square or Fisher's exact tests. | | | | | | |

| **Table S2. Performance of the three diagnostic models and sensitivity analyses in the validation sample (n=149).** | | | | | | | | | |
| --- | --- | --- | --- | --- | --- | --- | --- | --- | --- |
| **Model** | **M0** | | | **M1** | | | **M2** | | |
|  | **β_original_** | **β_shrunk_**  **(s=0.9249)** | **β_penalized_** | **β_original_** | **β_shrunk_**  **(s=0.9175)** | **β_penalized_** | **β_original_** | **β_shrunk_**  **(s=0.9193)** | **β_penalized_** |
| Age, 1-year increase | – | – | – | -0.050820 | -0.046625 | -0.048163 | -0.050992 | -0.046877 | -0.047870 |
| Fever, presence | 1.727861 | 1.598142 | 1.688091 | 1.586681 | 1.455731 | 1.531269 | 1.569582 | 1.442926 | 1.496003 |
| Oxygen saturation (room air), 1% increase | -0.136564 | -0.126312 | -0.128422 | -0.187662 | -0.172174 | -0.175829 | -0.244274 | -0.224563 | -0.225975 |
| Neutrophil count, 1×10^9^ cell/L increase | – | – | – | – | – | – | -0.207961 | -0.191180 | -0.193822 |
| Intercept | 12.013340 | 11.111438 | 11.266010 | 20.602260 | 18.901936 | 19.286960 | 27.423690 | 25.210763 | 25.403820 |
| **Discrimination** | | | | | | | | | |
| C-statistic (95% CI) | 0.817 (0.774–0.861) | 0.817 (0.774–0.861) | 0.819 (0.776–0.862) | 0.793 (0.747–0.840) | 0.793 (0.747–0.840) | 0.794 (0.747–0.841) | 0.819 (0.775–0.862) | 0.819 (0.775–0.862) | 0.819 (0.776–0.863) |
| **Calibration** | | | | | | | | | |
| Calibration-in-the-large (95% CI) | 0.032 (-0.217–0.282) | 0.032 (-0.217–0.282) | -0.030 (-0.221–0.280) | -0.318 (-0.587 to -0.048) | -0.317 (-0.587 to -0.048) | -0.292 (-0.559 to -0.026) | -0.372 (-0.656 to -0.089) | -0.372 (-0.656 to -0.089) | -0.374 (-0.658 to -0.091) |
| Slope  (95% CI) | 1.281 (1.030–1.531) | 1.385 (1.114–1.655) | 1.338 (1.077–1.598) | 0.874 (0.688–1.060) | 0.952 (0.749–1.156) | 0.923 (0.727–1.119) | 0.767 (0.608–0.927) | 0.835 (0.661–1.008 | 0.821 (0.651–0.991) |
| Overall miscalibration test^§^, p-value | 0.046 | 0.007 | 0.016 | 0.001 | 0.012 | 0.012 | <0.001 | <0.001 | <0.001 |
| Calibration accuracy test*, p-value | 0.011 | 0.002 | 0.004 | 0.045 | 0.131 | 0.188 | <0.001 | 0.001 | <0.001 |
| Brier score^ | 0.160 | 0.162 | 0.161 | 0.171 | 0.171 | 0.170 | 0.168 | 0.166 | 0.166 |
| McFadden's R^2^ | 0.399 | 0.391 | 0.397 | 0.318 | 0.329 | 0.331 | 0.327 | 0.347 | 0.344 |
| Cutpoint°:   - Sensitivity (95% CI) - Specificity (95% CI) | 0.5271  0.897 (0.852–0.931)  0.653 (0.575–0.725) | 0.5251  0.897 (0.852–0.931)  0.653 (0.575–0.725) | 0.5563  0.893 (0.848–0.928)  0.659 (0.581–0.730) | 0.6045  0.853 (0.803–0.894)  0.659 (0.581–0.730) | 0.5961  0.853 (0.803–0.894)  0.659 (0.581–0.730) | 0.5940  0.853 (0.803–0.894)  0.659 (0.581–0.730) | 0.6890  0.849 (0.799–0.891)  0.713 (0.638–0.780) | 0.6751  0.849 (0.799–0.891)  0.713 (0.638–0.780) | 0.6767  0.849 (0.799–0.891)  0.713 (0.638–0.780) |
| **DECISION CURVE ANALYSIS** | | | | | | | | | |
| Net reduction in intervention (%), threshold point:   - 0.1 - 0.2 - 0.4 - 0.6 - 0.8 | 0  4.5  17.3  19.8  26.5 | 0  4.5  17.3  19.8  26.2 | 0  4.5  17.3  19.8  26.2 | 1.4  8.1  14.3  20.3  25.0 | 1.4  7.9  14.3  20.2  25.6 | 0.5  7.2  14.3  20.2  25.7 | 1.4  6.2  13.8  19.9  27.4 | 0.5  5.7  13.4  20.1  26.9 | 0.7  5.0  13.6  20.1  27.1 |
| β, coefficient of the logistic regression from the development sample; CI, confidence interval; AUC, Area Under the Curve. ^§^Based on chi-square test with 2 degrees of freedom for testing unreliability (index U); *Based on the Spiegelhalter Z-test; ^The average squared difference in predicted and observed values; °Based on the Youden J empirical. | | | | | | | | | |
